# Lives and Livelihood, Not Quite a Trade-Off: A Cross-Country Analysis of the Short-Term Impact of COVID-19 Mortality on Real GDP

**DOI:** 10.1101/2023.02.13.23285835

**Authors:** Jing Lian Suah

**Affiliations:** Central Bank of Malaysia, Malaysia

**Keywords:** COVID-19, GDP, Two-Way Fixed Effects, Instrumental Variables, Trade-Off

## Abstract

**Introduction:** A supposed lives-livelihood trade-off (LLTO) has been at the centre stage of the COVID-19 pandemic, where policymakers often attempt to balance the health cost of COVID-19, including deaths, and the economic cost of lockdowns.

**Methodology:** This paper uses country-level panel (longitudinal) data on real GDP, stringency of non-pharmaceutical interventions (NPIs), economic policy support, COVID-19 deaths, and vaccination to quantify the short-run LLTO. Beyond descriptive analysis, adjustments were made — (1) two-stage least squares instrumental variables in a cross-sectional setting using pre-pandemic institutional quality as the excluded instrument, and (2) two-way fixed effects in a panel data setting.

**Findings:** Real GDP is negatively associated with COVID-19 deaths, as does more stringent containment measures. However, the offsetting positive association of real GDP with economic policy support is substantial. A historical decomposition of average real GDP that the positive attribution of fiscal support roughly equates the negative attribution of lockdown stringency and COVID-19 mortality.

**Conclusion:** Cross-country empirical evidence suggests no direct tradeoff between the economy, and public health. A change in policy thinking from a LLTO paradigm to a ‘no trade-off’ entails economic policy treating public health goals as invariant in supporting incomes through adequate, direct, and timely means.

## 1 Introduction

COVID-19 was, concurrently, a global public health, and a severe economic crisis. At its core was the presumed existence of a live-livelihood trade-off (LLTO), i.e., a determinate trade-off between economic well-being, or growth, and that of the effective control of the COVID-19 pandemic. This is unsurprising, as COVID-19 was unprecedented for many policy stakeholders. Rarely in modern history that crises placed multiple disciplines at primacy, especially that of both economics and epidemiology. For governments, this sets the stage for a tug-of-war, and conflict, between multiple schools of thoughts, and interest groups in the policy-setting process. The LLTO policy paradigm can be distilled essentially into the following lines of thinking — ‘is there an acceptable equilibrium level of COVID-19 deaths (or other severe outcomes of interest)?’, ‘how many people should die from COVID-19 to accommodate the economy?’, or ‘how much of economic growth can we sacrifice for the sake of controlling COVID-19?’. This paradigm draws parallels with a familiar problem in macroeconomics, the Phillips curve, which theorised a short-run trade-off between inflation and the output gap, unemployment, or real GDP. However, later renditions proposed there is no such trade-off in the long-run, or if the trade-off is exploited by attempting to control both variables ^1^. Recent research, however, finds the lack of such trade-off, and that the anchoring of inflation expectations was the principalis modus operandi (Hazell et al. 2022). In a similar tone, a higher-level take on the LLTO asks then if believing in a trade-off lands both the economy, and the state of the pandemic, in a ‘bad’ equilibrium (economy tanks and pandemic escalates), as opposed to a ‘good’ equilibrium (pandemic is contained, and economic cost is smaller) if policy actors were coordinated to prioritise public health goals. More formally, the policy function of the government in the non-LLTO paradigm entails treating public health goals as a constraint, rather than a variable. There are certainly limits as to what can be studied at this juncture, as the pandemic is still ongoing amid continuous emergence of new viral variants, and the first-generation vaccines had just been distributed widely, but bivalent adaptations are not yet available globally. Nonetheless, a retrospective evaluation of the short-run LLTO in the first two years of the pandemic is warranted, just as new disease outbreaks are emerging globally, e.g., monkeypox.

In the earliest days of the COVID-19 lockdowns in 1Q-2Q 2020, Kaplan et al. (2020) used a structural model (heterogenous agents neokeynesian model; HANK) calibrated to US data to illustrate a LLTO where higher deaths correspond to lower economic welfare cost. As more global data emerged, empirical studies of the LLTO were pursued. Ngo et al. (2022) used cross-country data in a random effects model, and a quantile panel data regression to argue that the LLTO is more prevalent where macroeconomic conditions are less favourable.

However, they noted the short study period spanning only June 2021, before the widespread introduction of vaccines, and the maturation of the Delta variant. Moreover, there could be methodological issues concerning endogeneity in the LLTO nexus arising from time-specific unobserved confounders, or simultaneity in policy levers. Decerf et al. (2021) used an adjusted ratio of number of poverty-years against life-years from 3 developed, and 3 developing economies up to June 2020 (the first COVID-19 wave) to posit an LLTO that differs according to state of economic development.

Empirical economic research also provides a view counter to the LLTO paradigm. Using cross-country data on COVID-19 and economic performance up to early 2021, Quah (2021) noted that NPIs may mitigate the negative economic effects of deaths, but weighs on economic activity, hence generating opposing effects. Guimbeau et al. (2020), using historical data spanning the “Spanish Flu” and World War I on Brazil, found that pandemic deaths had a negative effect on economic activity, even controlling for war deaths. The critique then is that NPIs may have been implemented to address the transmission of the influenza virus, and potentially had an effect on economic activity. Barro et al. (2020), drawing from cross-country evidence, arrived at a similar landing point. Correia et al. (1918) takes a longer term view in the US, but accounting explicitly for the variation in the implementation of NPIs. They found that states that implemented more decisive NPIs, and hence better health outcomes, experienced faster medium-term employment growth. Aum et al. (2021), however, posit that even in the absence of lockdown-type measures in South Korea, localised COVID-19 outbreaks adversely affected the labour market through reduced hiring by small firms, but with heterogeneity across sectors, skill levels, age and pay. Lin & Meissner (2020) further found that at the local-level, there is inadequate evidence that supports a link between job losses, mobility reductions from “stay-at-home” orders, and COVID-19 deaths at the local level. Rather, policy actions, and COVID-19 developments elsewhere generate “spillovers”, underscoring the importance of coordinated policy responses. This paper is most closely related to Islamaj & Mattoo (2021), who used cross-country data in a two-way fixed effects regression setup, and a IV regression, instrumenting the stringency of NPIs with lagged monthly industrial production growth. They argued, firstly, that an observed trade-off across countries is absent, and, secondly, its absence is policy emphasis on testing. However, like earlier cross-country studies, it suffers from the validity of the excluded instrument, and a short follow-up period in the data prior to the maturation of Delta, and global vaccination. Insofar as the health-economy nexus concerns an integrated health and economic policy strategy in containing COVID-19, the reaction of individuals to policies are certainly useful insights. Carrieri et al. (2020), in a randomised controlled trial in Italy, found that there is heterogeneity in the relative valuation given to policies that are framed as favouring health at the expense of the economy, or vice versa. More proximal exposure to the economic impact of COVID-19 tend to create a bias in the economic direction, while paternalistic framing on the health side creates a bias in the health direction. Relatedly, Settele & Shupe (2022), using online experimental data noted that a lower perceived economic of NPIs, or personal exposure to COVID-19-related health risks reduces personal responsiveness to such a trade-off.

Abstracting from the health-economy nexus, the COVID-19 pandemic certainly accompanied a notable economic downturn, as well as deaths in astronomic proportions. Cicala (2021) documents a sizeable increase in the incidence of extreme economic stress, indicated by households served utility severance notices due to a failure to pay for utilities, as well as demand for financial aid. Carletti et al. (2020), using firm-level data from Italy, documented a sizeable equity shortfall and drop in profits in light of the COVID-19 pandemic and subsequent lockdowns. Baker et al. (2020) documented a sizeable increase in economic uncertainty linked to the COVID-19 pandemic, and estimated a noticeable impact on aggregate economic performance. This general observation that the pandemic was associated with sizeable economic damage holds even when abstracting to historical pandemics. Jordá et al. (2020) found that real interest rates were persistently depressed decades after pandemics in Europe, juxtaposing the brief and meniscule impact of wars. From the health perspective, Mulligan (2021) documents the substantial rise in non-COVID excess deaths, including from substance abuse, as the pandemic escalated. However, health costs may arise from both the accompanying economic condition, and more directly the epidemiological state. Guo et al. (2020) zooms into data from China and found a significant effect on the incidence of deterioration in mental health conditions from both exposure to COVID-19 and the loss of livelihoods.

That there is a non-negligible economic cost associated with the pandemic gives rise to a plausibly important role for economic stimulus. Suah (2020) argues that the role of economic policy in a pandemic is to ensure that COVID-19 is contained, and to support income without compromising on health priorities. Atkeson et al. (2020) highlights the first of these principles, where economic policy that facilitates widespread testing, yields substantial economic benefits. Adherence to quarantines post-detection facilitate lower prevalence and deaths, as opposed to a low testing and detection situation, hence leading to higher business activity. An extension with vaccination further amplifies the economic gains by keeping deaths and prevalence low. Bell et al. (2021) motivates the second principle. Zooming into the UK employer-employee earnings data, the loss in earnings, and distributional impact across age, skill, sector, and unionisation, during the COVID-19 crisis were similar to past recessions. The alleviation of economic constraints, e.g., income, become crucial to the extent that they prevent compliance to NPIs, or make compliance less feasible. Alfaro et al. (2020) documents some of these concerns for emerging market economies (EMEs), especially where informal labour markets dominate. Moreover, they highlighted the primacy of policy in preserving formal labour matching arrangements. Casado et al. (2020) assessed that the rollout of fiscal stimulus in the US was crucial in boosting private consumption expenditure while ‘stay-at-home’ orders were implemented.

This paper ties together these elements systematically in an empirical analysis to characterise the nexus between COVID-19 deaths, NPIs, vaccination, real GDP growth, and economic policy support. Specifically, we ask (1) if cross-country data, and adjusted analyses, suggests a trade-off, and (2) to what extent does economic policy support play a role. We attempt these questions using cross-country national-level economic, health and policy variables in an instrumental variables (IV) linear regression approach using a cross-sectional setup, and a two-way fixed effects (TWFE) approach using a panel data setup. The subsequent sections explain the data, descriptive analysis, adjusted analysis, and finally conclude.

## 2 Data

The data has two parts — public health and the economy. Both data groups are compiled directly by CEIC data, from national authorities, the WHO, and respective data providers. These include OxCGRT (Hale et al. (2020)), and WEF (Porter et al. (2008), and Schwab (2019)). Data on the COVID-19 pandemic at the country-level are equivalently compiled by Roser et al. (2020) in an open-source database. For our analysis, we used the following variables — (1) the quarter-on-quarter growth of seasonally adjusted GDP in constant price local currency (real GDP) from national authorities, (2) the stringency index from OxCGRT, (3) daily new COVID-19 deaths per population from JHU CSSE, and national authorities, (4) the economic policy support index from OxCGRT, and (5) the institutions subindex from WEF’s Global Competitiveness Index, which excludes healthcare institutions. This study includes 55 countries with complete data throughout the 8 quarters, covering 34 AEs, and 21 EMEs, as classified by the International Monetary Fund (IMF),as shown in figure 1.

**Figure 1:**
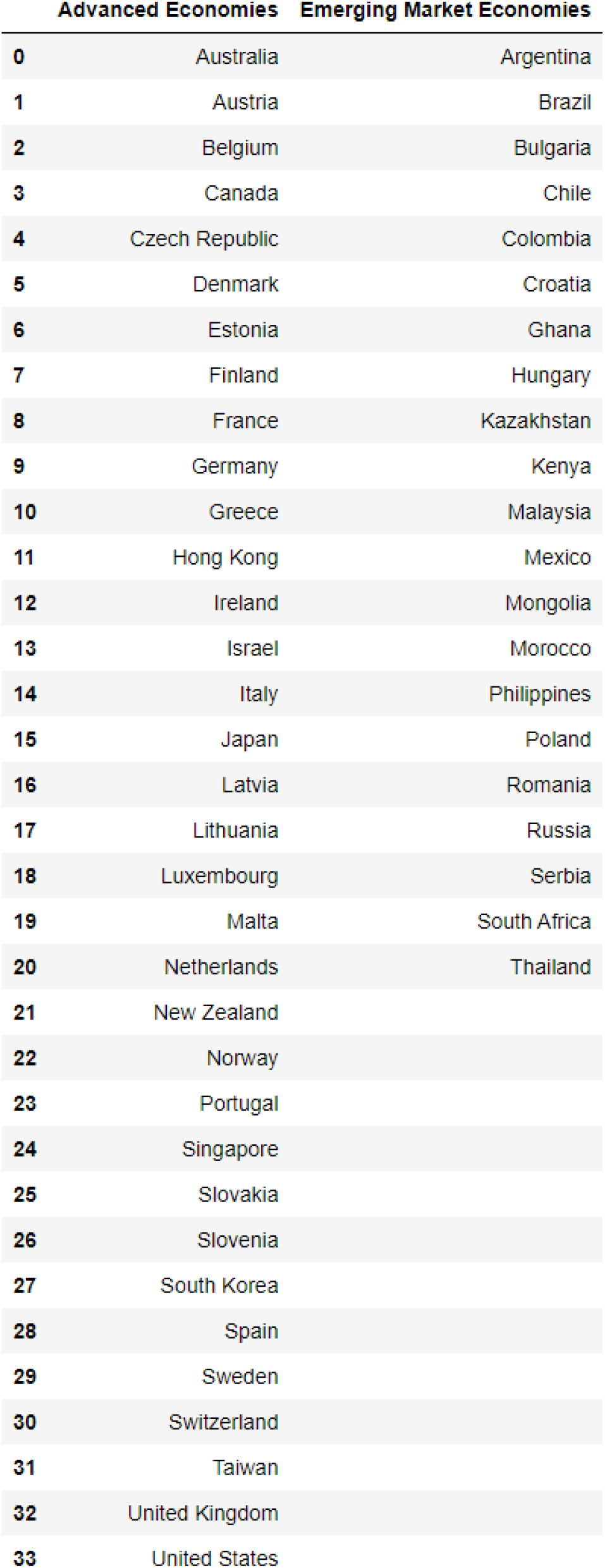
List of Countries Included in the Study by Country Classification

Some data undergoes transformation to facilitate interpretation. The daily series are transformed into quarterly frequency, with COVID-19 vaccinations per hundred population, and new COVID-19 deaths per thousand population as quarterly sums. The stringency, and economic policy support indices are transformed into quarterly averages. Population data is taken compiled by CEIC, and published by respective national authorities. The institutions index is available at an annual frequency, and all quarters in the same year takes the same value. However, only the 2019 value of the institutions index is included in the data set, as it is used as the excluded instrument for the cross-sectional analysis. The stringency, economic policy support and institutions indices are expressed on a 0 to 100 scale, as from OxCGRT. The analysis period, excluding the GCI institutions index, span 8 quarters from 1Q 2020 to 4Q 2021. This corresponds to the start of the pandemic, the first wave, emergence of pre-Omicron variants of concern (Alpha, Beta, Gamma, and Delta), the global vaccination drive, to the first Omicron subvariant (BA.1, and BA.2).

## 3 Descriptive Analysis

This section will narrate the data using two sets of scatter plots — (1) the cross-sectional (CX) representation of 8-quarter sum / averages on the left, and (2) a panel (longitudinal) representation of quarterly sum / averages on the right of respective figures. In the CX representation, each observation represents one country. In the panel representation, each observation represents one country for the corresponding quarter.

In figure 2, cross-country data on real GDP growth and the incidence rates of deaths are, in general, negatively correlated. However, this negative correlation may reflect a variety of endogeneity. For instance, a worse country-specific outbreak may trigger stronger NPIs, hence hurting the economy. Alternatively, countries with worse outbreaks may reflect a culmination of less responsive institutions, hence inadequate and / or untimely economic stimulus. This relationship holds weakly in both the CX and panel representations.

**Figure 2:**
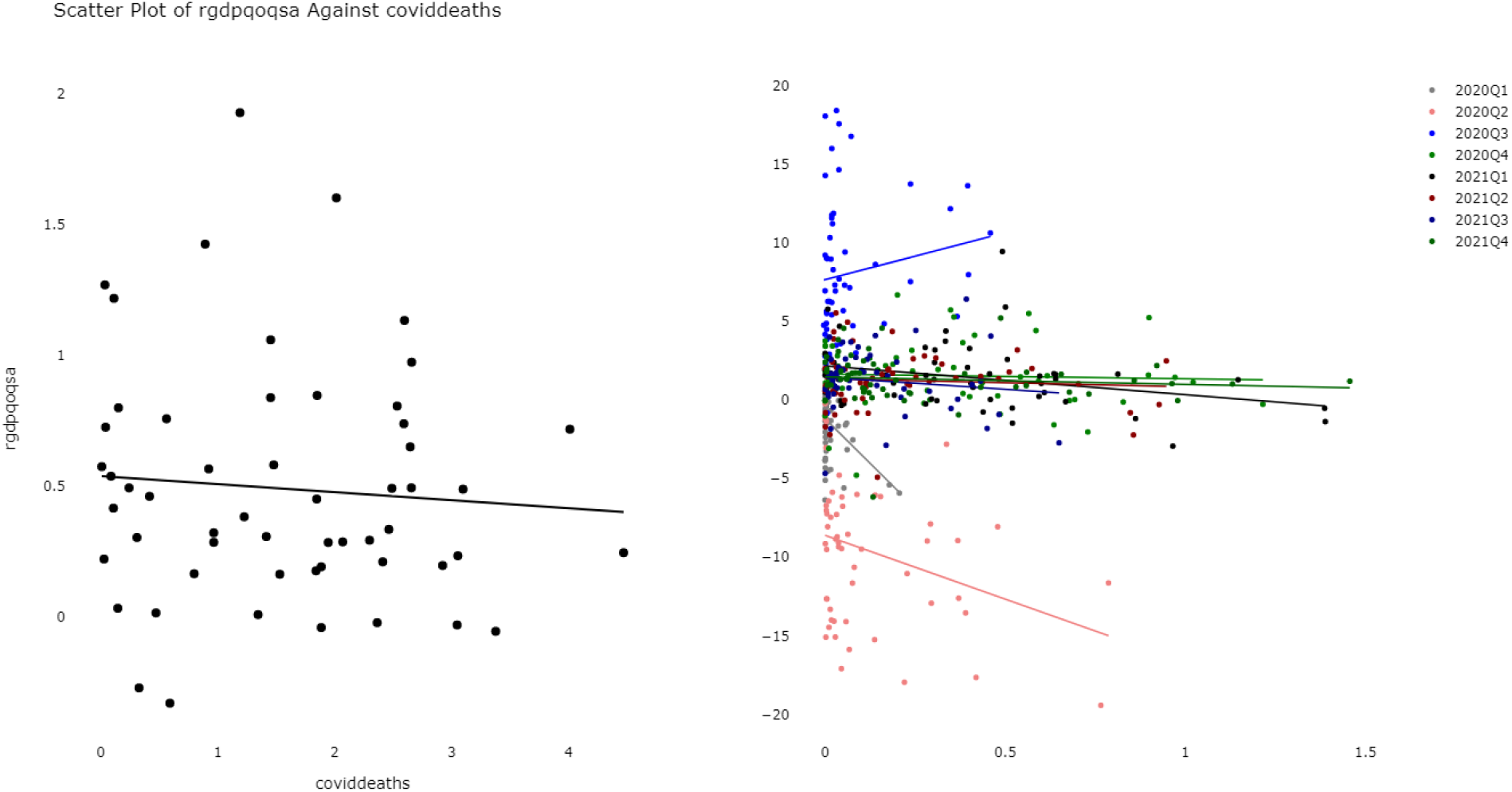
Scatter Plots of COVID-19 Deaths per Thousand Against Real GDP QoQSA Growth

Indeed, figure 3 shows that countries with higher COVID-19 deaths tend to have implemented stricter NPIs. A simple explanation is that NPIs are simply a response to the state of the outbreak. Additionally, countries with worse initial macroeconomic states may hesitate on implementing NPIs, and eventually forced to escalate to even stricter NPIs, once the outbreak proves to be difficult-than-initially-expected to contain, supporting the narrative in Decerf et al. (2021). While the CX representation suggests a relatively weak relationship, the panel representation suggests otherwise. This competing illustration reflects that the shifting willingness to implement NPIs, or mortality-NPI link, over time. NPIs appeared stricter earlier in the pandemic than later in the pandemic, before the mortality-NPI link completely breaks down by 4Q 2021 when Omicron hit, and when booster doses were rolled out in some countries (Roser et al. 2020).

**Figure 3:**
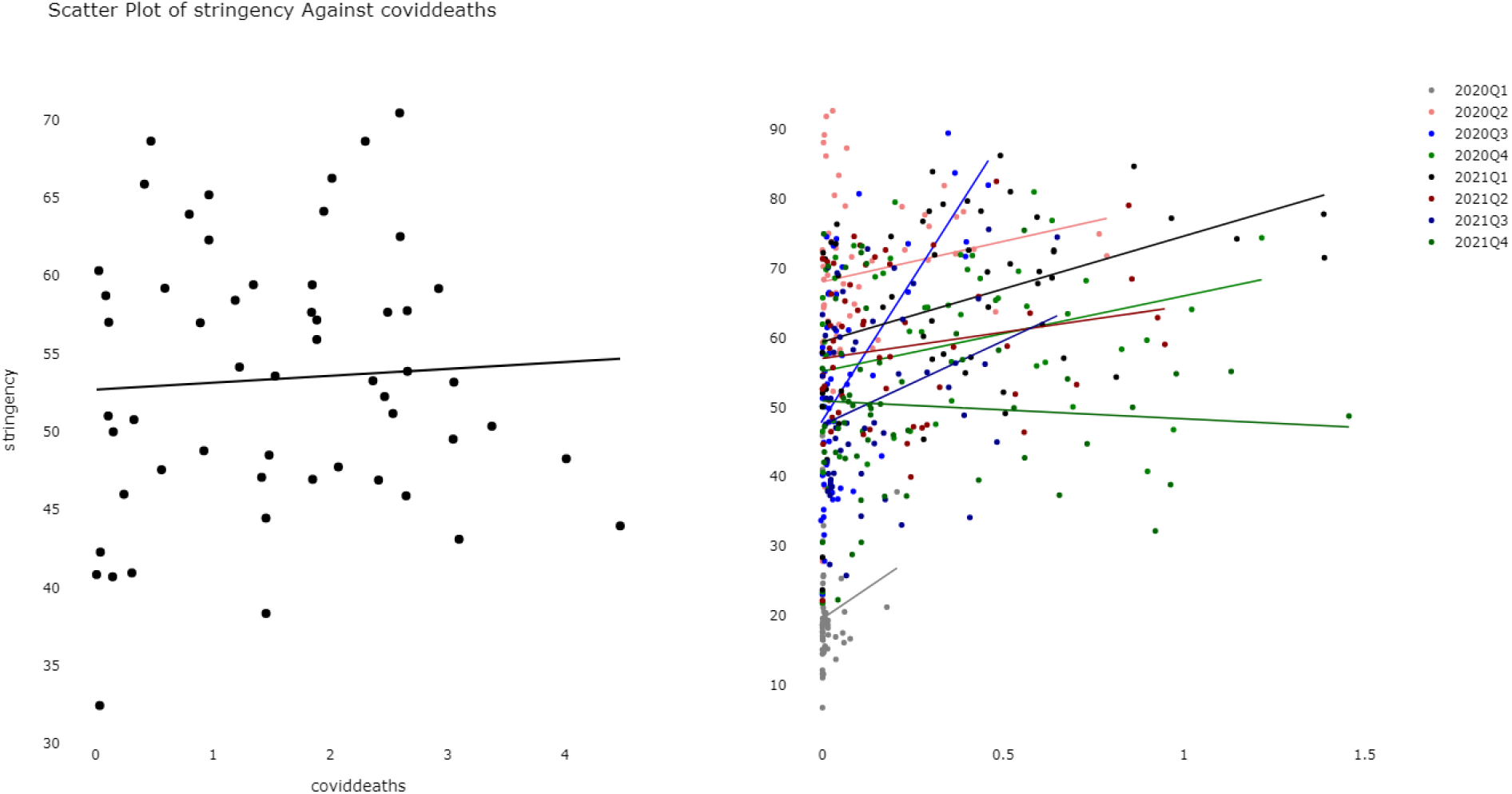
Scatter Plots of COVID-19 Deaths per Thousand Against Stringency Index

Unsurprisingly, figure 4 shows that countries with stricter lockdowns experienced weaker economic growth during most parts of the pandemic. This observation lends support to the view in Quah (2021), amongst others, that lockdowns may yield a direct economic cost. However, whether this negative association arises from concurrent spikes in COVID- 19 mortality rates, the supply and demand impact of lockdowns, or a mixture of both is uncertain.

**Figure 4:**
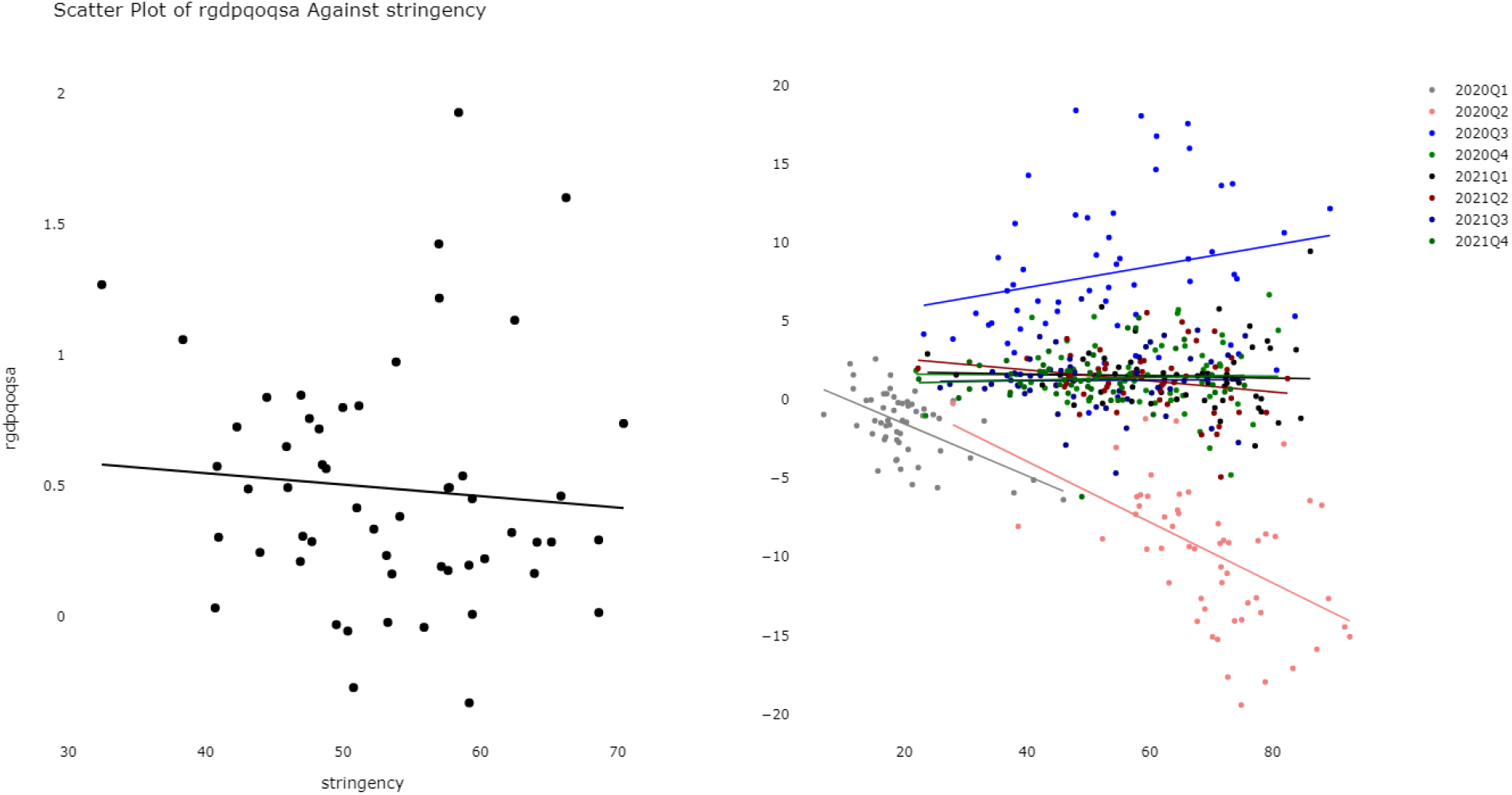
Scatter Plots of Stringency Index Against Real GDP QoQSA Growth

However, figure 5 suggests that not all countries with strict(er) NPIs implemented large(r) economic stimulus packages. This may reflect cross-country differences in fiscal policy space, responsiveness of policy platforms, and institutional capacities. The panel representation unveils that the NPI-stimulus link may be changing over time as the state of the pandemic evolved, such as when new viral variants become global, when vaccines were introduced, or simply when there is a lull in the transmission of COVID-19.

**Figure 5:**
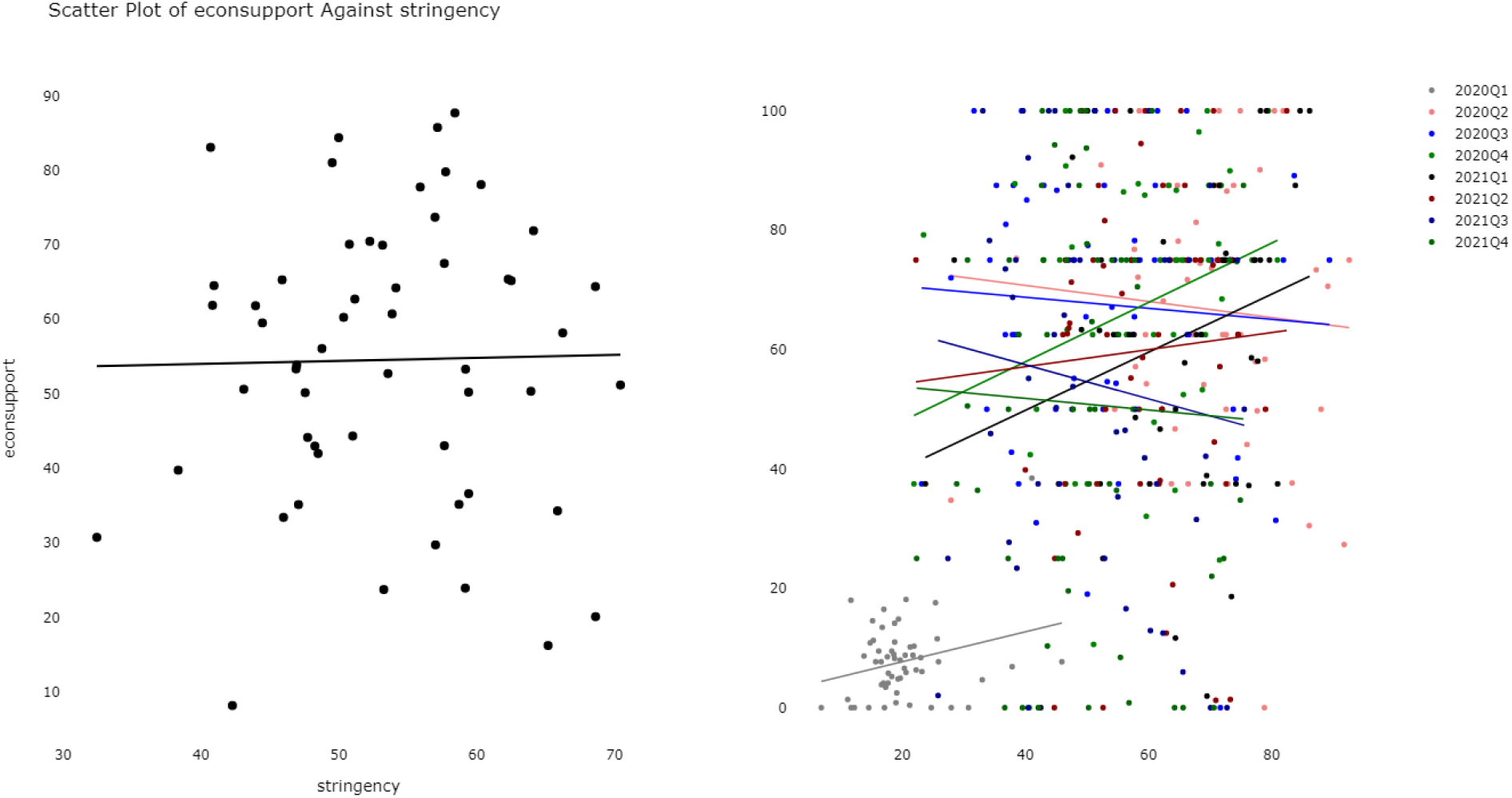
Scatter Plots of Stringency Index Against Economic Policy Support Index

Due to the multitude of confounding that may have underpinned the observed data described above, an adjusted analysis is warranted before the associations in the LLTO nexus can be established.

## 4 Adjusted Analysis

### 4.1 Methodology

We pursue two empirical strategies — (1) a a two-staged least squares (2SLS) instrumental variables (IV) approach, and (2) a two-way fixed effects (TWFE) estimator. Both methodologies are complementary.

#### 4.1.1 Two-Staged Least Squares Instrumental Variables

To estimate the CX associations in the LLTO nexus, we use a 2SLS IV approach. A simple OLS is also estimated as benchmark. In this setup, all variables are collapsed into their 8-quarter average / sum by country, as described in the data section, effectively forgoing the panel data structure. Country-specific confounders that are time-invariant still need to be controlled for. Economic policy support is a response variable, which ultimately may determines the extent to which NPIs can be implemented, hence confounds the “trade-off”. Economic policy space, in turn, can be explained by largely time-invariant factors such as institutions, state capacity, or fiscal ideology. The short timespan (2020-21) used in the analysis further supports that these factors may be time-invariant. Hence, the institutions index from 2019 is used as the excluded instrument for economic policy support to account for confounding in the LLTO nexus. In essence, only the variation in stimulus measures that is associated with pre-pandemic institutional quality, which by time sequence is exogenous, is used in the analysis.

The 2SLS IV uses fitted values of the instrumented variable from the first stage regression, in lieu of the observed values, to estimate the LLTO equation in the second stage. The instrumented variable (economic policy support) is *X*_*i*_. The included instruments 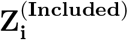 act as covariates in both the first and second stages, comprising stringency, and COVID-19 deaths. In the second stage equation, 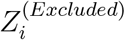 is the excluded instrument (GCI institutions index from 2019). Finally, the coefficient on the instrumented variable *β* is interpreted as the variation in real GDP that is explained by only the exogenous variation in economic policy.

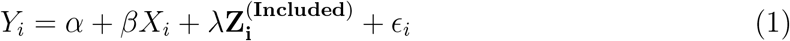

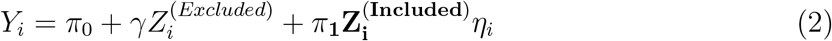

Identification of *β* rests on (1) relevance, where there is a non-zero correlation between institutions and economic policy, and (2) exclusion restriction, where institutions affect GDP only through economic policy support. The first condition can be confirmed through the first stage regressions. As a heuristic, we will report the first-stage F-statistic. Moreover, state capacity influences the ability of governments to garner economic resources, e.g., through capital markets and its network of public institutions, to spend during the epidemic. Stronger institutions also determines the readiness and resilience of channels for the government to funnel resources to afflicted households and firms, e.g., through pre-existing welfare programmes, developmental credit facilities, public outreach, as well as the degree of elite capture within the bureaucracy. The GCI institutions index covers 8 broad areas, of which none are directly related to public health — security, social capital, checks-and-balances, public sector performance (regulation, legal and electronic framework), transparency, property rights, corporate governments, and the government’s future orientation. To the extent that the index measures only the business, and economic aspect of institutional qualities, the exclusion restriction holds. These are documented in the 2019 version of Porter et al. (2008) in Schwab (2019).

#### 4.1.2 Two-Way Fixed Effects

The TWFE regression setup includes country-specific fixed effects *α*_*i*_, and quarter (time)- specific fixed effects *α*_*t*_, for countries *i* and quarters *t. Y*_*it*_ is the explained variable — real GDP QoQSA growth. The vector of coefficients *β* on the vector of variables of interest and covariates **X**_**it**_ is estimated with the “within” estimator. **X**_**it**_ contains the remaining variables that vary between and within countries — COVID-19 deaths, stringency, and economic policy support.

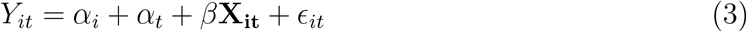

This method accounts for quarter-specific and country-specific unobservables through the fixed effects, which may confound the link between real GDP, stringency, and economic policy support, and estimates the average within-country associations in the LLTO nexus. For instance, quality of institutions, and state capacity to manage the epidemic may matter. These are potentially “long-term” factors, i.e., time-invariant over the study period, and are country-specific. Moreover, there may be common shocks over the course of the pandemic, e.g., the emergence of new viral variants that exhibit signs of vaccine escape. These are common factors across all countries, but unique to specific time frames.

### 4.2 Findings

#### 4.2.1 Two-Staged Least Squares Instrumental Variables

Figure 6 shows the coefficient estimates of the IV 2SLS regression, and the OLS version as reference. In both versions, the signs are identical, but differ quantitatively. The 95% confidence bounds are similarly displayed, together with the p-values. The coefficient estimates are imprecise, which may reflect either the sample size constraint of 55 economies, or that the LLTO paradigm is indeed not supported by CX empirics.

**Figure 6:**
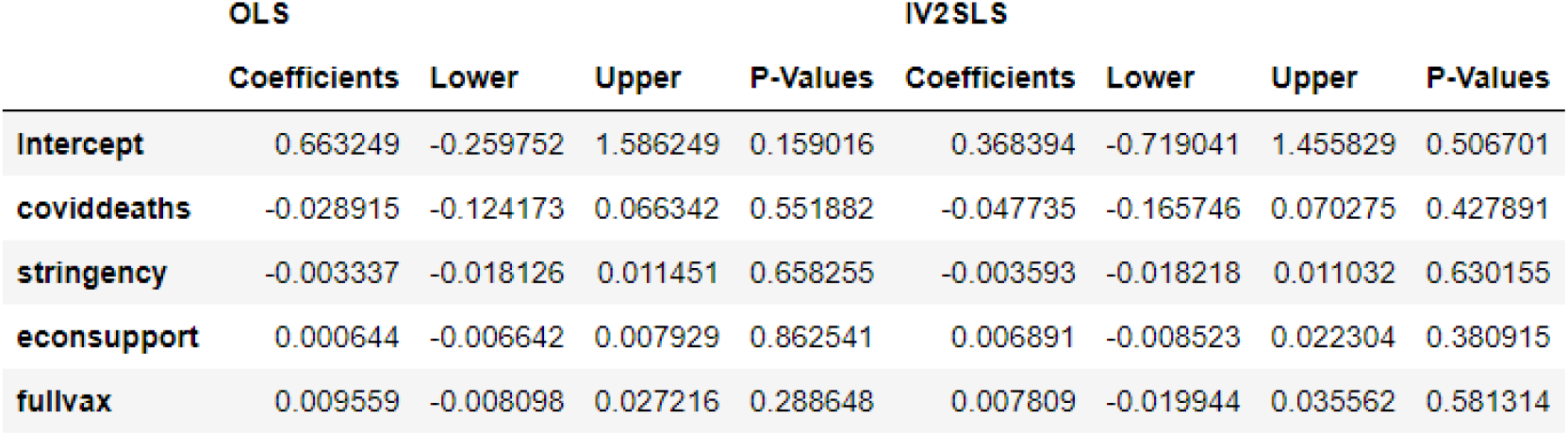
IV 2SLS Coefficient Estimates

Figure 7 further shows the diagnostics, including the first stage F-statistic for the IV 2SLS, which assists in interpreting if the relevance assumption (that pre-pandemic institutional quality has a non-zero association with economic policy support) holds.

**Figure 7:**
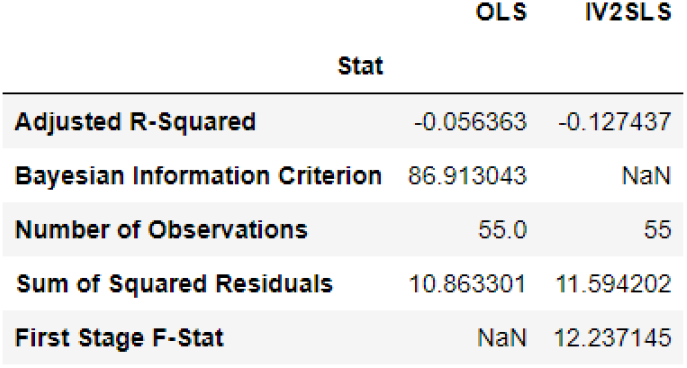
IV 2SLS Diagnostic Statistics

As the raw coefficients are hard to interpret in context of real GDP growth, and that the relative importance of each variable in their associations with real GDP growth may be unclear, figure 8 shows the historical decomposition of average real GDP QoQSA growth across all countries in the CX analysis, but excluding the intercept, and residual terms. On average, subject to statistical imprecision, stringency contributed to about -0.2 percentage point (ppt) of real GDP growth, and COVID-19 mortality rates to about -0.1 ppt. This is more than offset by economic policy support, which contributed about +0.4 ppt to real GDP growth, suggesting a major role for economic policy support, and that stimulus implemented throughout the pandemic was adequate. Vaccination, in and of itself, contributed a small +0.01 ppt.

**Figure 8:**
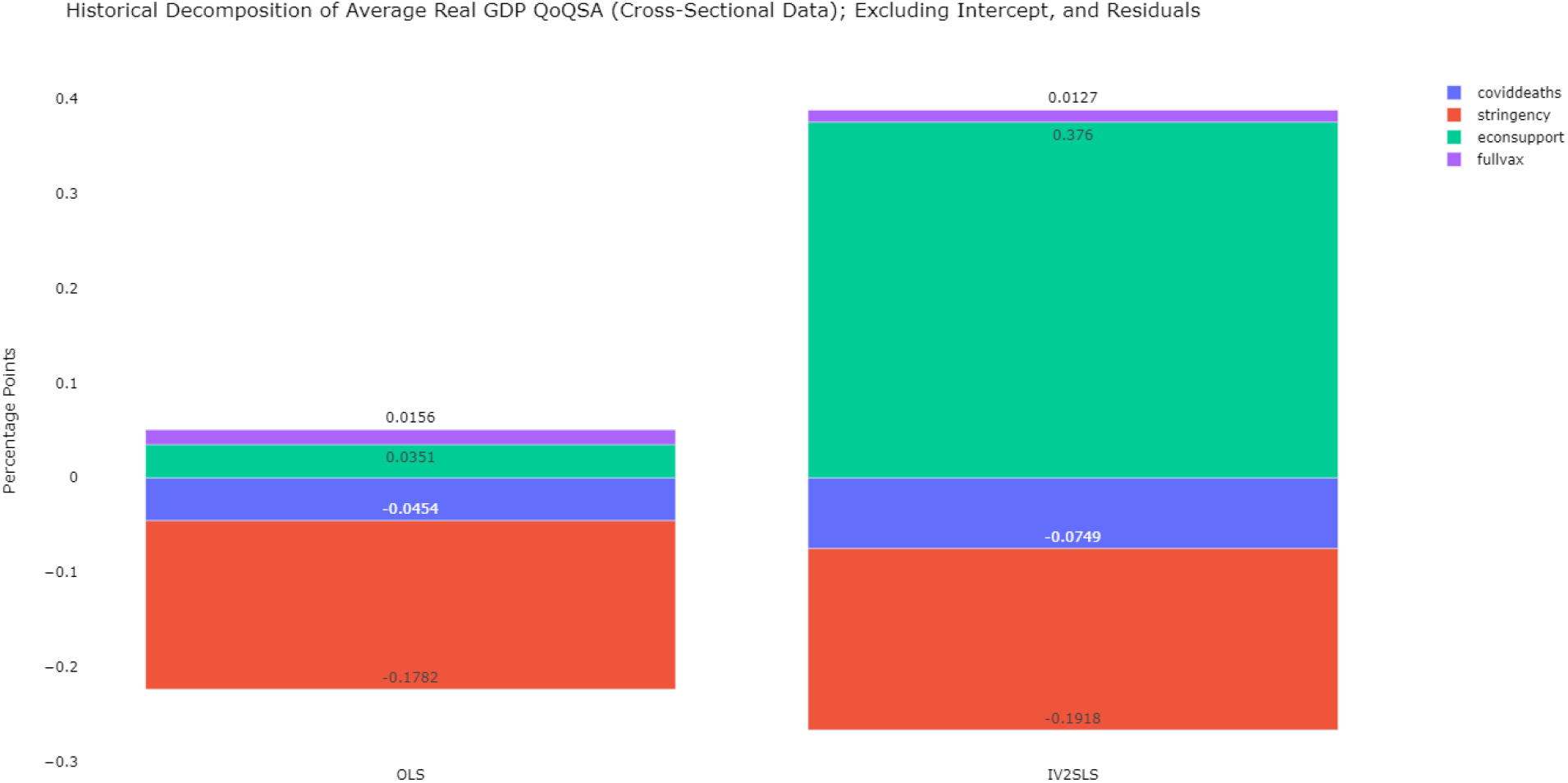
IV 2SLS Historical Decomposition of Average Real GDP QoQSA Growth

#### 4.2.2 Two-Way Fixed Effects

Figure 9 shows the TWFE regression estimates. The pooled version, and the country-only fixed effects version are included as benchmark. Unlike the CX analysis, both the signs and quantum of the coefficient estimates differ between all three specifications. In the TWFE model, only the negative association between COVID-19 mortality incidence rate and real GDP growth is precisely estimated at the significance level of 10%. Reasons for imprecision are likely the same as in the CX analysis, a combination of short follow-up period, sample size constraints due to aggregation, and / or that the LLTO nexus is intrinsically unclear. Figure 10 shows the diagnostic statistics.

**Figure 9:**
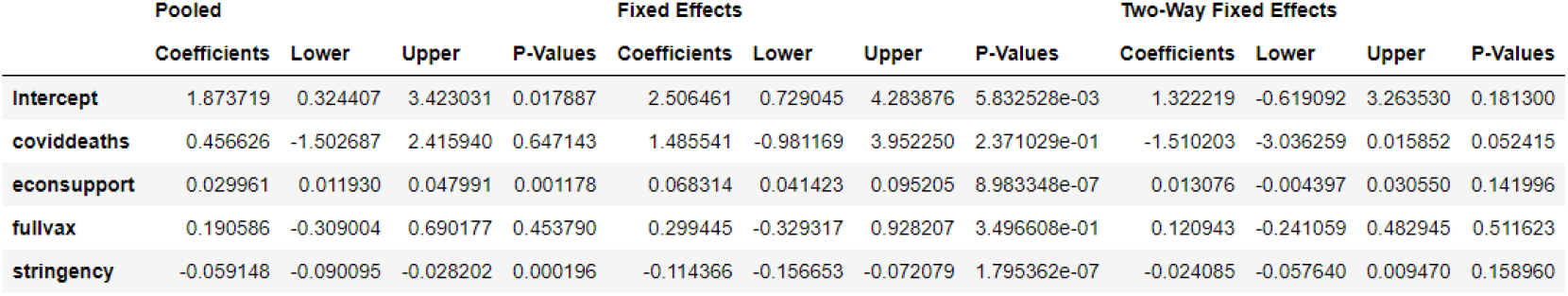
TWFE Coefficient Estimates

**Figure 10:**
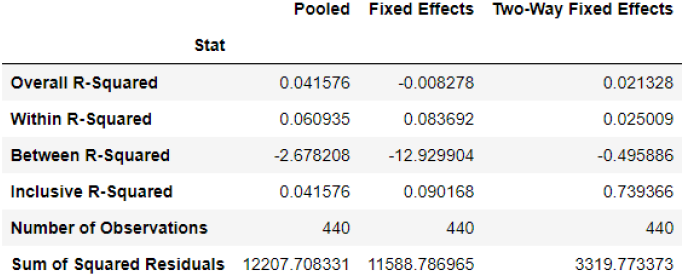
TWFE Diagnostic Statistics

**Figure 11:**
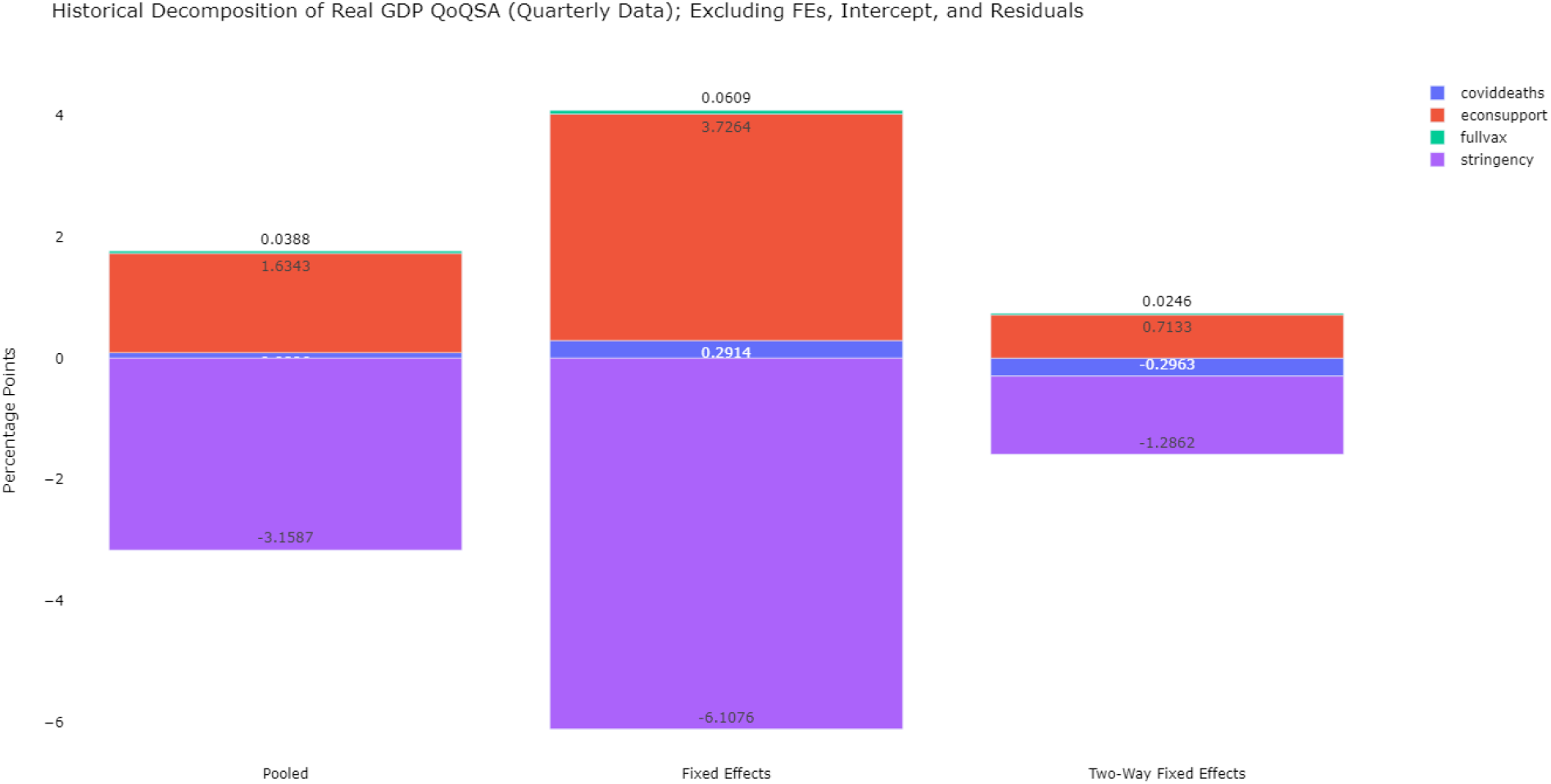
TWFE Historical Decomposition of Average Real GDP QoQSA Growth

Figure 10 then shows the historical decomposition of observed average real GDP growth in terms of COVID-19 mortality, economic policy support, vaccination, and stringency of NPIs. The contributions are generally smaller in the TWFE than the FE and pooled setups, reflecting the importance of controlling for time-specific, and country-specific unobservables, e.g., institutional quality, emergence of new variants, and shifts in policy thinking. Quantitatively, the TWFE estimates differ from the IV 2SLS. The contribution of NPIs is far larger at -1.3 ppt, and death rates slightly larger at -0.3 ppt. The contribution of vaccination is larger by about 2-fold at +0.025 ppt. However, economic stimulus now far from offsets the contributions from NPIs and COVID-19 mortality at +0.7 ppt. While this supports that there is a primal role for economic policy in the pandemic, the observed implementation in 2020-21 may be inadequate, subject to estimation imprecision.

## 5 Discussion

This study offers two empirical observations. Firstly, a descriptive analysis of COVID-19 mortality, stringency of NPIs, economic stimulus, and economic growth, in both CX, and panel representations. Secondly, an adjusted analysis of the LLTO nexus, including vaccination, using IV 2SLS (CX), and TWFE (panel) regressions. The analysis suggests that there is no direct trade-off between the economy and public health. A worse pandemic and more stringent NPIs both hurt economic growth independently, subject to estimation imprecisions. In the TWFE, the weakly significant negative association between COVID-19 mortality, and real GDP growth, while other variables are statistically insignificant lends further support to the view that the LLTO paradigm was mistaken. Similarly, the IV 2SLS analysis also found imprecise estimates in the same direction. While the closely related Islamaj & Mattoo (2021) found a lack of trade-off, even when a shorter study duration, excluding the main segment of the global vaccination drive, the analysis accepted the existence of a trade-off that was simply weakened by testing. In a LLTO paradigm, the expected sign of the association between COVID-19 mortality, and real GDP, should either be positive, or a precisely estimated zero, coupled with a negative contribution from NPIs. The empirics appear at odds.

This finding has important policy implications, albeit more relevant for the ongoing monkeypox pandemic, and future public health crises. Policy should, first and foremost, prevent escalating transmission and deaths. Economic activity stands to be damaged further if deaths escalate, which tend to trigger even more stringent NPIs, doubling down on the economic cost of an outbreak. Furthermore, vaccination has an important role to play in epidemics or pandemics of vaccine-preventable diseases by weakening the observed infection-mortality link. Moreover, from a purely economic point of view, vaccination is associated with positive dividends to economic growth. Importantly, economic policy during public health crises ought to be adequate, timely, and direct to support incomes, so that public health goals, be it vaccination, containment, mitigation, or a combination of both, can be achieved in full (Suah 2020).

However, the analysis does have some limitations. Firstly, the study looks only at the short-term associations. A long-term characterisation of the LLTO ought to consider the impact of widespread ‘long COVID’, lifetime all-cause mortality as population health deteriorates due to repeated COVID-19 outbreaks, and productivity spillovers from COVID-19 related disruptions in the global population. Moreover, causality cannot necessarily be inferred in this type of observational studies.

Finally, to close the discussion, the policy message of the paper can be distilled into the following — that the world ought to move from a ‘lives-livelihood trade-off paradigm’ to a ‘no trade-off paradigm’. An conceptual illustration of both paradigms are in figure 12.

**Figure 12:**
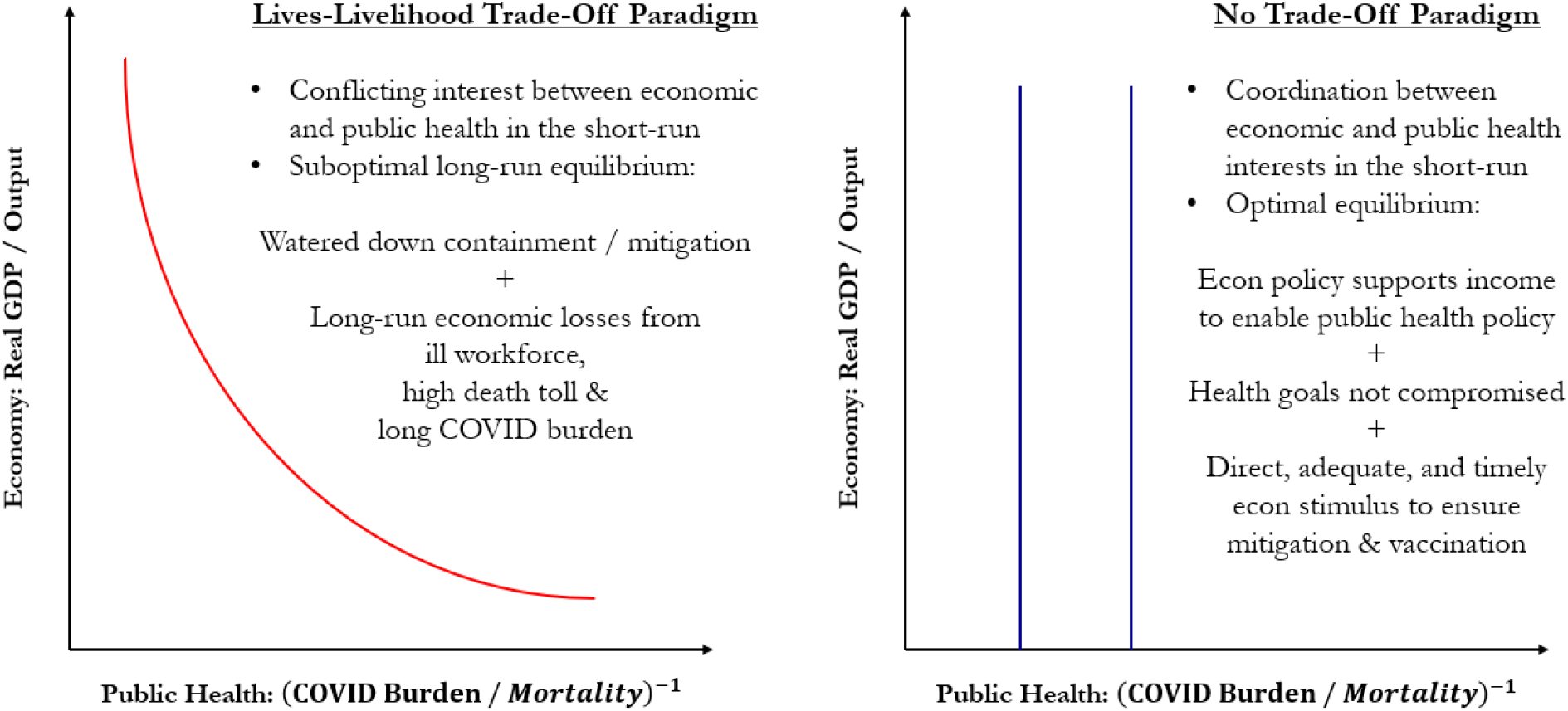
Conceptual Relationship Between Economy and Public Health

In the LLTO paradigm, economic and public health interests are conflicting, reflective of most of policymaking throughout the pandemic in 2020-22. This lands the world in a suboptimal equilibrium, suppose that multiple equilibria exists, where a combination of watered down but prolonged pandemic control measures, and further economic losses from COVID-19 mortality, an unhealthy workforce, and higher all-cause mortality hurt both the states of the economy, and the public health. In the Phillips Curve analogy, this corresponds to a downward sloping short-run LLTO, but a flat / vertical long-run LLTO, arising from time- inconsistency in policymaking institutions.

The suboptimal equilibrium, in this line of thinking, can be avoided or escaped through coordination, and commitment devices. Specifically, policy should treat public health goals as the constraint, rather than a variable, in a similar fashion that most central banks became inflation-targeters to stave inflation, rather than to exploit the implied trade-off between inflation, and output / unemployment. In this paradigm, economic policy supports income to enable public health policy through direct, timely, and adequate stimulus.

## 6 Conclusion

Cross-country empirical evidence suggests no direct tradeoff between the economy, and public health. A change in policy thinking from a LLTO paradigm to a ‘no trade-off’ entails economic policy treating public health goals as invariant in supporting incomes through adequate, direct, and timely means.

## Data Availability

This research uses only publicly available aggregated data, and does not involve any human participants. All scripts and data vintages required for replication are available at https://github.com/suahjl/llto-macro.

## Ethical consideration

As this research uses only publicly available aggregated data, and does not involve any human participants, institutional review board (IRB) approval is not required.

## Data and replication statement

This research uses only publicly available aggregated data, and does not involve any human participants. All scripts and data vintages required for replication are available at **github.com/suahjl/llto-macro**.

## Acknowledgement

This paper has benefited from the following individuals / groups for their critical review and feedback.

**•** Boon Hwa Tng, Central Bank of Malaysia, Malaysia

**•** Thevesh Thevananthan, Central Bank of Malaysia, Malaysia

**•** Seminar participants at the 16^th^ Vaccine Congress

## Footnotes

1 Most standard macroeconomics textbook, at various levels of technical involvement, would have sufficiently covered this debate. For instance, see Mankiw (2020).

